# Zero-Dose and Dropout Children in Nigeria: A Comparative Machine Learning Analysis of Childhood Immunization Cascade Failure Using the 2023–24 Nigeria Demographic and Health Survey

**DOI:** 10.64898/2026.09.05.26362351

**Authors:** Eloghosa Nosa-Ihaza, Michael Chiebuka Ogidi, Odeke Stephen Okello, Uyioghosa Nosa-Ihaza

## Abstract

**Background:** There are two distinct failure points in childhood immunization: children who receive no immunization (“zero-dose”); and children who start but do not complete the immunization series (“dropout”). Two failure points to address are possible: ‘zero-dose’, where children receive no first doses; and ‘dropout’, where children begin but do not complete the immunization series. These are plausibly different failure points that require different interventions. Nigeria has the largest number of children with zero doses in absolute numbers in the world. The newly released Nigeria Demographic and Health Survey (NDHS) 2023–24 is used to describe failure cases and to compare the performance of machine learning techniques with standard regression in predicting them.

**Methods:** We analyzed 4,937 children aged 12-23 months (the age range covered by the WHO/UNICEF children’s recode) in the NDHS. Survey-weighted logistic regression was used to model ZDS and dropout (Penta1 received vs. Penta3 not received) and to compare them with the WHO/UNICEF national coverage estimates (WUENIC). With the same held-out test data, each of three additional algorithms - elastic net, random forest, and XGBoost - was compared to logistic regression, and sampling weights were added to each. Geographic clustering, after adjusting for individual-level effects, was quantified using a mixed-effects model with a state-level random intercept.

**Results:** Zero-dose prevalence was 37.3% (implied Penta1 coverage 62.7%), compared with the WUENIC national estimate of 71% Penta1 coverage; incomplete series coverage was at 9.1% (14.5% of Penta1 recipients), and full vaccination coverage was at 53.6%, which is lower than the administratively weighted global coverage estimates (67% Penta3). The associations of zero-dose status were strong and independent for maternal education, facility delivery, attending ANC, and household wealth, whereas these factors were surprisingly weak, albeit statistically significant for Maternal education and Facility delivery, and not significant for the other factors in the dropout group. After full adjustment, religious affiliation (among Muslims) remained an independent predictor of zero dose, even though clustering by state was adjusted for. Survey-weighted logistic regression performed best for both outcomes (zero-dose AUC=0.825; dropout AUC=0.613), whereas XGBoost performed worst for both. Only 4.7% of the residual variation in dropout could be accounted for by state of residence, but 11.2% could be accounted for by zero-dose status.

**Conclusions:** Zero-dose status as well as dropout are characterized by very different predictor profiles and are, therefore, separate policy problems and should not be treated as a single “coverage gap”. There was no improvement in prediction performance in terms of algorithmic complexity compared to a correctly specified logistic regression for either outcome.

## 1. Introduction

Immunization remains one of the most cost-effective public health interventions for children, yet coverage has not reached high levels, with gaps across countries. These gaps can be better understood by dividing immunization gaps into two separate failures: (1) never starting the immunization schedule, often referred to as “zero-dose” children, defined by the WHO/UNICEF as children who have received no dose of the diphtheria-tetanus-pertussis-containing vaccine (WHO, 2020); and (2) children who start the schedule but fail to complete it, usually measured by attrition between the first and third doses of the pentavalent (DTP-HepB-Hib) vaccine. They are NOT the same problem. A child who drops out after the first dose has passed the threshold – by definition, has never been zero-dose – the threshold is not about the first dose; it is about continuing to be in contact with the health system. The two failure points reasonably represent two separate types of failure - the first is the failure to generate demand, and the second is the failure to deliver continuity of care and follow-up systems - and to address them as a single undifferentiated “coverage gap” could lead to resources being used on the wrong lever.

Nigeria is a case study of particular importance for exploring this division. The number of zero-dose children in the country is the highest in absolute numbers of any country in the world, with estimates by WHO/UNICEF currently exceeding two million in recent reporting cycles. There is also a clear north-south and religious split in coverage, which is well documented and has been described as associated with socioeconomic disadvantage, access to facilities and, for some in the northeast, longstanding insecurity which hinders routine outreach. The 2023–24 Nigeria Demographic and Health Survey (NDHS) was conducted by the National Population Commission between December 2023 and May 2024, and was supported by the ICF/DHS Program; it is the first nationally representative, sub-nationally disaggregated survey since the last DHS round in 2018, and provides updated empirical evidence on the distribution of these two failure points.

There are direct antecedents in previous quantitative studies on immunization coverage in Nigeria, but two studies are very similar in design. Drawing on the Nigeria DHS conducted in 2018, Aheto et al. (2022) applied such Bayesian multilevel models in the same age group (12-23 month) as included in this study to examine the receipt of Penta1 and Penta3-given-Penta1 in 2018, with maternal education, vitamin A receipt, and health-card ownership being significant correlates of both outcomes. In this study, using a different method and the most recent available data, Sato (2023) analyzed five rounds of DHS from 2003–2018, and also revealed that the salience of maternal education and household wealth, as explained by this study, was very different across the two outcome types, again independently finding that maternal education and household wealth were important factors in explaining the downward trends of zero-dose and under-immunization rates, but less important in explaining the downward trends of dropout rates. A distinct line of research has focused on the use of machine learning for classification of the zero-dose population; earlier, Biswas, Tucker and Bauhoff (2023) tested predictive algorithms for the zero-dose population for three countries – India, Mali and Nigeria – but did not test for dropout nor compare different algorithms using the same held-out data. To best present this new study, the finding should not be seen as the introduction of an untested analytical approach, but rather as extending this closest previous work in three ways: its application of a four-algorithm, its proper survey-weighted machine learning comparison of these algorithms in tandem to both the zero-dose and dropout categories, and its testing, rather than assuming, that algorithmic complexity is directly predictive of either outcome; it was deliberately chosen to not include the substantive indicator of health-card ownership because the card is typically issued only when a child goes into the vaccination system, and is thus too close to the outcome definition itself to be an independent predictor.

A methodological question arises from the adoption of this particular toolkit of analytical tools. It is commonly believed that machine learning techniques, such as gradient boosting, will outperform simpler regression techniques on structured survey data, though this is not always true. The logistic regression model performed better than an XGBoost model and a random forest model on a similarly structured cohort of sociodemographic and clinical predictors of uncontrolled hypertension among adults in 2025 analyses of the National Health and Nutrition Survey (ENSANUT) in Mexico. Whether a similar pattern is observed for a structurally similar prediction problem in a different domain and setting- sociodemographic prediction of a binary health outcome from cross-sectional survey data- is not an inevitability, but rather a testable one with direct implications for future research on immunization in Nigeria.

This study seeks to fulfill these gaps by having four specific aims. First, national and sub-national disaggregated estimates of zero-dose status and Penta1-to-Penta3 dropout are estimated for Nigerian children aged 12-23 months, the age band commonly used in reporting coverage in WHO/UNICEF surveys, and then compared with the corresponding WHO/UNICEF WUENIC coverage figures. Second, we undertook a survey-weighted logistic regression analysis to determine the individual (person) and household (clusters of individuals) factors associated with each outcome, testing whether either or both failure points were proximately associated with distinct factors. Third, we extend this comparison to three other algorithms that are appropriately weighted for the complex survey design - elastic net, random forest, and XGBoost - and test all four on the same held-out data to directly test whether algorithmic complexity is better for either outcome. Fourth, we estimated the proportion of the residual variance in each of the outcomes that is explained by geography, while controlling for individual- and household-level predictors, using multilevel logistic models with a random intercept at the state level. These analyses are designed to provide not only new national-level estimates but also a methodologically informed assessment of the difference, if any, between the factors associated with never initiating immunization and those associated with not completing it.

## 2. Literature Review

### 2.1 Global and Regional Epidemiology of Zero-Dose and Dropout

Nigeria’s role in this global context is out of balance (Causey et al., 2021). The country has also consistently been ranked with the highest absolute numbers of zero-dose children in the country compared with any other country in the world, and in a regional analysis of the WHO African Region, Nigeria was one of the four most populous countries with more than 80% of zero-dose children between 2019 and 2022 (Mboussou et al., 2024). The same analysis revealed a trend relevant to this study’s benchmark comparison: In 19 of the 47 African countries reported, administrative coverage was higher than the WUENIC blended estimate, which was a result of data quality issues in routine immunization rather than survey error, thus supporting this study’s interpretation that it is a known tendency of the benchmark comparison, not a flaw in the survey methodology.

### 2.2 Determinants of Immunization Gaps in Nigeria

There is already a large descriptive and inferential literature that has tested correlations between incomplete immunization in Nigeria and maternal education, household wealth, place of delivery, attendance at antenatal care, and geographic zone, which this study’s results mostly replicate. Jean Baptiste and colleagues (2024) reported that the geospatial model of the Nigeria MICS/NICS 2021 showed that children in weak and vulnerable socio-economic groups were more likely to be zero dose and under-immunized, and that there were positive relationships between the percentage of zero dose children and child stunting and low maternal literacy (Birhanu et al., 2023). A recent household survey across six states confirmed this study’s finding that attendance to antenatal care and delivery in a health facility were the most protective against zero-dose status, as the rates among children with fewer than four ANC visits (or at home) were significantly higher at 35% in each of the six states surveyed, while the rates among children delivered in a health facility or by a skilled attendant were much lower.

There are two previous studies of a similar design to this one, which are convenient to compare directly with it. This study replicates the results of Sato (2023); it used five rounds of Nigeria DHS data (2003–2018), and applied Blinder-Oaxaca decomposition to explore trends in the prevalence of zero-dose, under-immunization, and dropout status, uncovering that although maternal education and household wealth are strong predictors of the observed declines in zero-dose and under-immunization status over time, both factors are less salient in predicting dropout status. Aheto and colleagues (2022) are even closer: They applied the 2018 Nigeria DHS and applied Bayesian multilevel models to the same 12-23 month age band used here and found that ownership of a health card or document, receipt of vitamin A, and maternal education level were significant for both Penta1 receipt and Penta3-given-Penta1 completion. In this prior study, the authors used card ownership as a substantive predictor; in this study, they deliberately used card retention as an independent predictor, and they found that this was a more appropriate approach because a card is not usually issued more than once after a child enters the vaccination system, resulting in circularity.

### 2.3 Machine Learning Approaches to Immunization Prediction

There have been relatively few studies that have conducted systematic comparisons of multiple machine learning approaches for immunization outcomes, particularly in Nigeria. The closest parallel is by Biswas, Tucker and Bauhoff (2023), who evaluated the performance of predictive algorithms for zero-dose risk classification from previous DHS rounds in India, Mali and Nigeria, but only for zero-dose classification and did not compare the performance of different algorithms using the same held-out samples and datasets. In other contexts, outside of Nigeria, a very similar four-algorithm approach was applied to Ethiopian vaccination data (Endehabtu et al., 2026) and resulted in gradient boosting and random forest classifiers outperforming other algorithms in terms of discriminative classification performance, with the best performing reaching an AUC of almost 0.97 among children aged 12–35 months, which is a much higher ceiling than found in this study, most likely because of differences in the richness of the predictor sets used (e.g., also including perception-based and service-utilization variables that were not included in typical DHS recodes), and not a contradiction of the core finding of this study about algorithmic complexity.

### 2.4 Algorithmic Complexity and Predictive Performance

This study’s main methodological result - that the survey-weighted logistic regression on both outcomes was the best performing among the three ensemble and boosting methods, elastic net, random forest, and XGBoost - is not without precedent, but contrasts with the popular notion that the two ensemble and boosting methods are always superior on structured tabular data. In another study, the logistic regression, random forest and XGBoost models analyzed on a similar set of sociodemographic and clinical variables yielded best performance for the random forest model (AUC = 0.75), followed by the logistic regression model (AUC = 0.61), and the XGBoost model (AUC = 0.54) for the diagnosis of uncontrolled hypertension in ENSANUT 2025 (Mendoza-Cano et al., 2025). The pattern that holds true in both of the studies is not as broad as a simple “simplicity wins” statement: while an ensemble method was superior to logistic regression in the Mexican study, this study’s results indicated that logistic regression was superior to all three machine learning alternatives, with the exception of the XGBoost model, which proved to be the worst. The common thread in both scenarios is that XGBoost was the poorest performer in every scenario, indicating a more specific methodological lesson than the field default: the benefits of gradient boosting for structured survey data in which there are a moderate number of sociodemographic predictors are not universal and should be assessed empirically and not assumed across domains and settings.

### 2.5 Summary and Study Rationale

As a whole, the body of literature in this study’s scope has a significant foundation, particularly in the work of Aheto et al. (2022) and Sato (2023), which illustrate the Nigeria-specific two-outcome comparison. The contribution of this study is therefore not in providing an unprecedented analytical approach but in making the first use of a four-algorithm machine learning comparison to both zero-dose and dropout together, with the new 2023–24 NDHS round, combined with a state-level multilevel robustness check and explicit benchmarking against current WUENIC estimates, thus extending rather than replacing the closest earlier work on this exact question.

## 3. Methods

### 3.1 Data Source and Study Population

Data for this study are drawn from the Nigeria Demographic and Health Survey (NDHS) conducted in 2023–24 by the National Population Commission and Federal Ministry of Health and Social Welfare with the technical assistance of ICF and the DHS Program, which was conducted between 1 December 2023 and 7 May 2024 (NPC, FMoHSW, and ICF, 2025). The survey was based on a stratified two-stage cluster design in which 1,400 enumeration areas (EAs) were sampled as primary sampling units (PSUs) across the 36 States and the Federal Capital Territory (FCT) of Nigeria, and within each EA, households were sampled. Analyses are based on the Children’s Recode (KR) file, which records each child born to an interviewed woman in the five years before the survey, and are combined with maternal and household characteristics from the Individual and Household Recode files.

The KR file’s vaccination history module — the source of both outcome variables in this study — is administered only to living children born in the three years preceding the survey (age 0–35 months at interview), consistent with standard DHS questionnaire design. This base population of 14,916 children was further restricted to children aged 12-23 months at the time of interview (n=4,937), the age band reported in WHO/UNICEF coverage reporting and in other previous studies in Nigeria (e.g., Aheto et al., 2022). There are two factors in this restriction. Second, it does not introduce downward bias in vaccination estimates because it excludes children who were too young to receive the last dose; in a diagnostic comparison of three-month bands of age, there was no evidence of this bias, with the 37.3–45.4% of children with zero doses across the age bands being roughly the same size as the 12–23 month band. Second, it produces zero-dose and dropout estimates directly comparable to WHO/UNICEF WUENIC national coverage figures, which report DTP1 and DTP3 coverage — Nigeria’s operational equivalent of Penta1 and Penta3 in the country’s combined pentavalent (DTP-HepB-Hib) schedule — using the same age convention. Results using the fuller 0–35-month sample are reported separately for robustness comparison.

### 3.2 Outcome Definitions

A two-stage cascade framework was used to create two outcomes. Gap A (zero-dose status) was defined as children who were coded as having received the pentavalent vaccine (confirmed from a vaccination card seen by the interviewer, reported by mother without a card being seen, or reported by mother on a card without a date) were considered as having received Penta1, while those who were coded as not received (including “don’t know” responses) were considered as zero-dose. Conventionally, a DHS coverage table “don’t know” is coded as “non-receipt,” which yields a prevalence estimate of 37.0%. However, coding this “don’t know” as “missing” rather than “non-receipt” (as would be done for other measures) and recoding the “unknown” as “missing” (affecting 23 of 4,937 children, 0.47%) yielded a materially similar result (37.0% versus 37.3%) indicating that the coding choice of “don’t know” as “non-receipt” is not sensitive to the outcome.

For children who received Penta1, the analogous variable used for defining Gap B (dropout) was the receipt of Penta3; those who were not administered Penta3 were considered as dropout, and those who received all three doses were considered as completed.

### 3.3 Predictor Variables

The following were selected as predictors: maternal education (categorized into four groups: none, primary, secondary, higher); household wealth quintile (Rutstein et al., 2004); urban/rural residence; geopolitical zone (grouped into six categories, following the convention used in the DHS final report); child’s religion (grouped into four categories: Hausa, Yoruba, Igbo, Fulani, in accordance with the DHS final report), child’s ethnicity (collapsed into four groups: Hausa, Yoruba, Igbo, Fulani, with all other non-missing, non-“other” codes combined into a single “other” category); child’s place of delivery (recoded to a binary indicator for facility versus home, following the standard DHS convention that codes 11 and 12 indicate home delivery, while other codes indicate a public or private health facility); maternal attendance of 4 or more ANC visits (in accordance with the DHS final report convention that codes 11 and 12 represent 4 or more visits); presence of self-reported barriers to accessing health care (four items were asked in this survey round: permission to seek care, cost, distance, and reluctance to attend alone — the four items fielded in this survey round of the standard access-barriers battery, birth order, child sex, and regular media exposure (a binary indicator for at least weekly exposure to newspapers, radio, or television, constructed from the three standard DHS media-frequency items).

Although health-card retention (whether the mother had the child’s vaccination card at the time of the analysis) was included in previous analyses (Aheto et al., 2022), it was omitted from the final set of predictors. This variable was found to show a very high association with zero-dose status (odds ratio 0.04) and also to induce a high discrimination value (AUC 0.80 to 0.90), which is the hallmark of data leakage: Possession of a vaccination card marks the first time when a child was vaccinated by the health care system, and this is mechanically related to the outcome and not an independent predictor.

### 3.4 Statistical Analysis

Survey weighting, primary sampling units (clusters) and sampling strata were used in all analyses and Stata’s survey (svy) estimation commands were used. Design-based (Rao-Scott-corrected) chi-squared tests were used to evaluate the association of each predictor with each outcome (Rao et al., 1948). Multivariable associations were calculated using survey weighted logistic regression and are reported as survey weighted adjusted odds ratios (aORs) and 95% confidence intervals (CIs).

Three other algorithms were fitted, all implemented in Python (scikit-learn and XGBoost libraries) (Pedregosa et al., 2011), and passed observation-level sampling weights directly to each algorithm’s sample_weight argument: elastic net regularized logistic regression (Zou & Hastie, 2005), random forest (Breiman, 2001), and gradient-boosted trees (XGBoost) (Chen & Guestrin, 2016). The 80/20 train-test split was generated once in Stata, stratified by Gap A outcome, and fixed with a documented random seed; thus, the same data were held out for prediction across all four algorithms and both outcomes. The discriminative performance was presented as a weighted area under the receiver operating characteristic curve (AUC) on the held-out test set.

To estimate the effect of geographic clustering in addition to the fixed effects already specified, random intercepts for state (37 levels: 36 states and Federal Capital Territory) were added to each of the two logistic regression models, and the residual intraclass correlation coefficient was computed to provide a measure of the proportion of the variation in the outcome that was not explained by the individual and household-level effects.

The estimates were compared to the national coverage estimates in the WHO/UNICEF Estimates of National Immunization Coverage (WUENIC) for the same age group for 2024.

## 4. Results

### 4.1 Sample Characteristics and Flow

In 2023-24, there were 27,783 children recorded in the NDHS Children’s Recode, of whom 14,916 were living children born in the preceding three years and therefore eligible for inclusion in the vaccination history module. Further restricting to the age band at interview (12–23 months) as per the WHO/UNICEF coverage age band resulted in a primary analytic sample of 4,937 children. The Gap A (zero-dose) model included 4,681 children (94.8% of all age-eligible) after data for one predictor were missing from the dataset. Of the 3,164 children in this sample with Penta1 data, 2,983 children (94.3%) had full predictor data and were included in the Gap B (dropout) model. A summary of this sample selection process is given in Figure 1 for each restriction stage.

**Figure 1.**
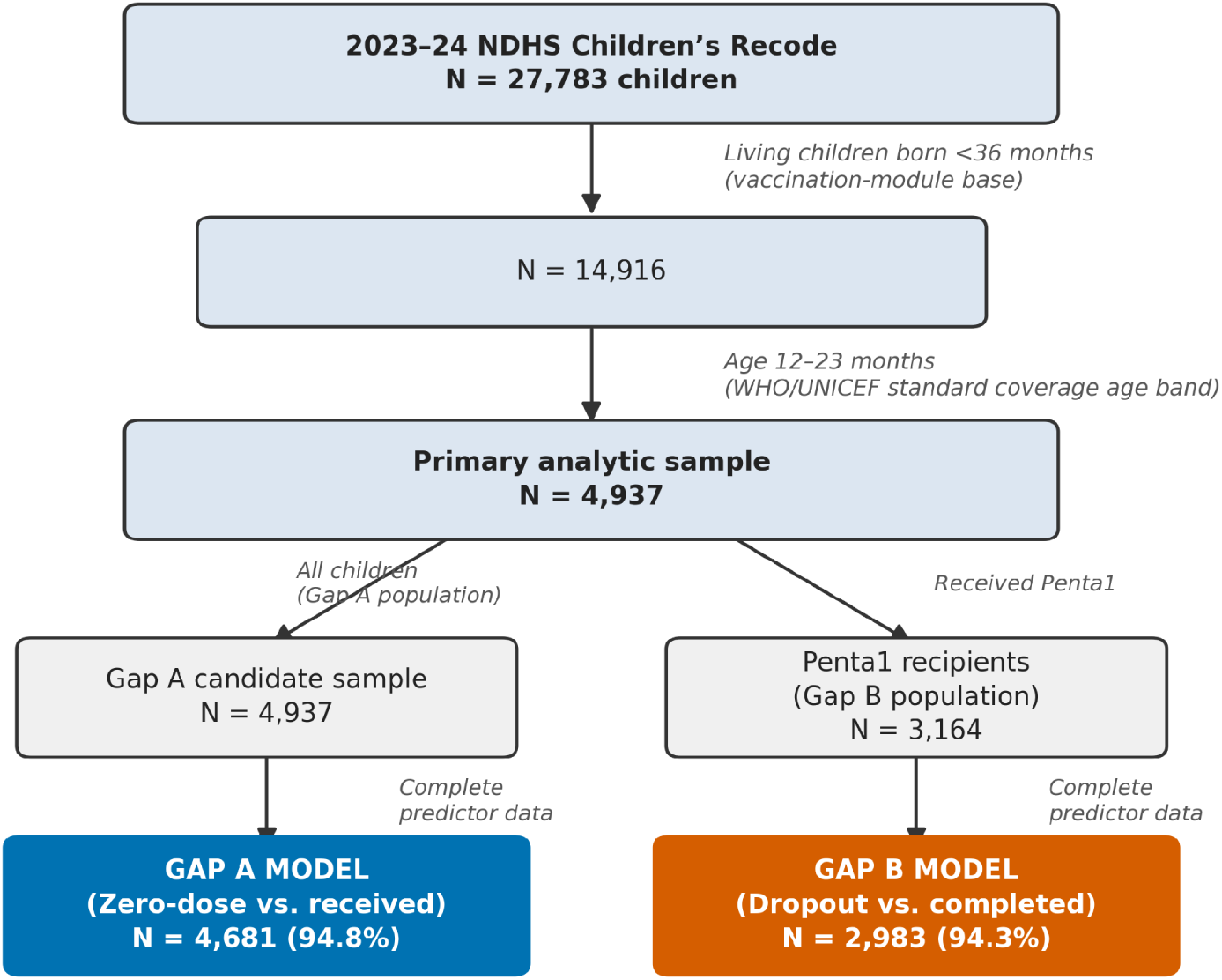
Sample selection flow diagram, from the full NDHS Children’s Recode to the final Gap A and Gap B model samples.

The weighted characteristics of the primary analytic sample are summarized in the table below (Table 1). The majority of the sample (60.1%) was in rural areas, with the North West zone having the highest proportion of children (38.1%) and the North East zone the lowest (16.0%); the three southern areas combined 26.1%. Maternal educational attainment was low; 44.0% reported no formal education. The distribution of wealth quintiles was fairly balanced (16.4–23.7% each quintile). The majority of religions reported (67.7%) was Islam, as is typical of the north. Four in 10 (44.6%) children were born in a health facility, and 56.7% of mothers reported four or more visits to the health care system during their pregnancy. There was very low health insurance coverage (2.6%).

**Table 1.** Weighted characteristics of the primary analytic sample (children 12–23 months, n = 4,937) Note: all estimates weighted using the normalized DHS sampling weight and adjusted for the complex survey design (74 strata, 1,286 PSUs).

| **Characteristic** | **% or Mean (SE)** |
| --- | --- |
| Residence: Urban | 39.9% |
| Residence: Rural | 60.1% |
| Zone: North West | 38.1% |
| Zone: North East | 19.6% |
| Zone: North Central | 16.0% |
| Zone: South East | 7.3% |
| Zone: South South | 8.3% |
| Zone: South West | 10.8% |
| Maternal education: None | 44.0% |
| Maternal education: Primary | 11.8% |
| Maternal education: Secondary | 33.5% |
| Maternal education: Higher | 10.8% |
| Ethnicity: Hausa | 39.0% |
| Ethnicity: Igbo | 9.9% |
| Ethnicity: Fulani | 9.0% |
| Ethnicity: Yoruba | 8.9% |
| Ethnicity: Other | 33.3% |
| Wealth: Poorest | 23.7% |
| Wealth: Poorer | 22.2% |
| Wealth: Middle | 18.9% |
| Wealth: Richer | 18.9% |
| Wealth: Richest | 16.4% |
| Religion: Islam | 67.7% |
| Religion: Other Christian | 25.1% |
| Religion: Catholic | 6.9% |
| Religion: Traditionalist | 0.4% |
| Religion: Other | 0.02% |
| Health insurance: No | 97.4% |
| Health insurance: Yes | 2.6% |
| Place of delivery: Home | 55.4% |
| Place of delivery: Facility | 44.6% |
| ANC 4+ visits: No | 43.3% |
| ANC 4+ visits: Yes | 56.7% |
| Regular media exposure: No | 64.2% |
| Regular media exposure: Yes | 35.8% |
| Child sex: Male | 50.9% |
| Child sex: Female | 49.1% |
| Birth order, mean (SE) | 3.68 (0.045) |
| Access barrier count (0–4), mean (SE) | 1.01 (0.027) |

### 4.2 Prevalence of Zero-Dose Status, Dropout, and Completion

The primary sample estimate showed that 37.3% received no dose (no Penta1), 53.6% were fully vaccinated (three doses of Penta1), and 9.1% dropped out (received only Penta1) (Table 2). In the coverage terms of WUENIC, that is, the percentage of children covered by a specific number of vaccines, it translates to 62.7% implied Penta1 (DTP1) coverage and 53.6% implied Penta3 (DTP3) coverage, which are both significantly lower than the corresponding 2024 national estimates for Nigeria of 71% (Penta1) and 67% (Penta3). The gap is consistent with a documented trend of higher administrative coverage (which contributes to the WUENIC blended estimate) than coverage based on household surveys in most African countries, and does not signal inconsistency in the present analysis.

**Table 2.**
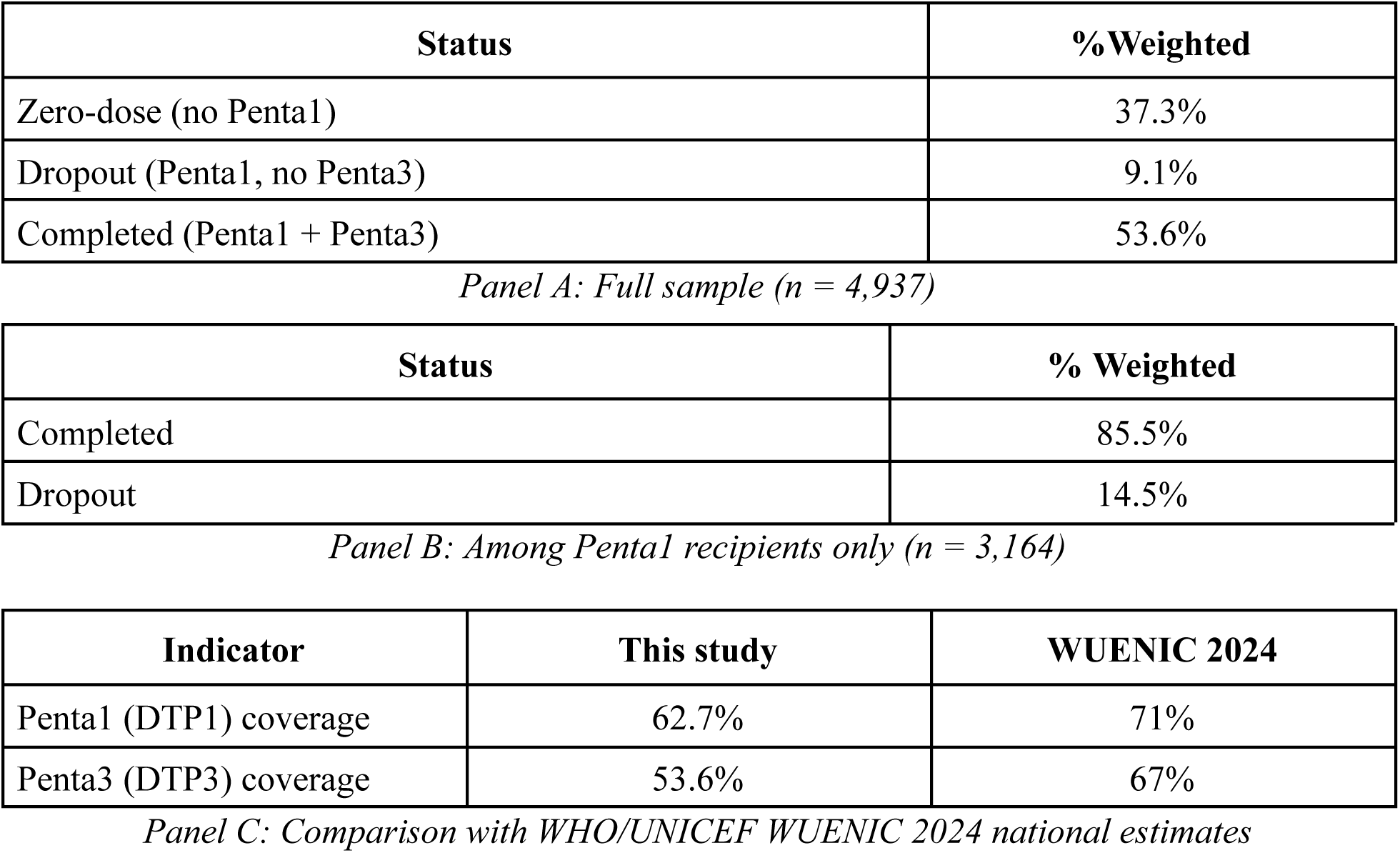
Immunization cascade status, children 12–23 months. Note: survey-based estimates below the WUENIC blended estimate are consistent with a documented pattern (Mboussou et al., 2024) in which administratively reported coverage, which contributes to WUENIC, exceeds household-survey-based coverage in a majority of African countries.

A diagnostic comparison across three-month age bands confirmed the age-restriction approach, with zero-dose prevalence being highest in the 0–11 months age band and the 12–23 months age band showing the lowest prevalence, and the 24–35 months age band having the highest prevalence (design-based F(1.96, 2531.77) = 24.56, p < 0.001).

A sensitivity analysis that re-coded “don’t know” vaccination responses as missing (rather than non-received) did not result in a significantly different zero-dose estimate (37.0% versus the headline 37.3%), but did only change the estimate of 23 of 4,937 children (0.47%), supporting the robustness of the headline outcome definition.

### 4.3 Bivariate Associations

All predictors except child sex were significantly related to zero dose in the bivariate analysis (Table 3). The highest sub-distinction was by maternal education (59.3% had no education compared to 5.3% for higher education; F(2,90, 3514,40) = 178,01, p < 0,001), facility delivery (54.0% for mothers who delivered at home compared to 16.7% for facility deliveries; F(1,1209) = 399.11, p < 0.001) and number of ANC visits (61.2% for mothers who attended fewer than 4 ANC visits compared to 19.6% for those who attended ≥ 4 ANC visits; F(1,1201) = 509,02, p < 0,001). There was also a significant difference in the prevalence of zero doses by geopolitical zone (7.5% in the South East to 53.9% in the North West) and by religious affiliation (13.7% among Catholics to 47.6% among Muslims).

**Table 3.** Bivariate association between predictors and Gap A / Gap B outcomes (weighted %, design-based F-test)

| Predictor / Category | % Zero-dose (A) | % Dropout (B) |
| --- | --- | --- |
| Residence: Urban | 22.5% | 15.4% |
| Residence: Rural | 47.2% | 13.6% |
| F, p | $F(1,1212)=139.18, p<.001$ | $F(1,1070)=1.23, p=.267$ |
| Zone: North West | 53.9% | 13.7% |
| Zone: North East | 36.0% | 16.8% |
| Zone: North Central | 40.0% | 19.0% |
| Zone: South East | 7.5% | 14.1% |
| Zone: South South | 12.8% | 8.5% |
| Zone: South West | 16.2% | 12.9% |
| F, p | $F(4.35,5271)=61.49, p<.001$ | $F(4.85,5194)=3.00, p=.011$ |
| Education: None | 59.3% | 17.1% |
| Education: Primary | 36.8% | 17.3% |
| Education: Secondary | 18.9% | 15.0% |
| Education: Higher | 5.3% | 6.4% |
| F, p | F(2.90,3514)=178.01, p<.001 | F(2.97,3182)=7.56, p<.001 |
| Ethnicity: Fulani | 55.2% | 17.9% |
| Ethnicity: Hausa | 51.5% | 13.3% |
| Ethnicity: Igbo | 7.0% | 11.9% |
| Ethnicity: Yoruba | 15.1% | 14.5% |
| Ethnicity: Other | 30.8% | 15.8% |
| F, p | F(3.60,4367)=77.13, p<.001 | F(3.94,4211)=1.14, p=.337 |
| Wealth: Poorest | 56.9% | 16.7% |
| Wealth: Poorer | 52.1% | 19.6% |
| Wealth: Middle | 36.2% | 16.1% |
| Wealth: Richer | 22.9% | 14.3% |
| Wealth: Richest | 6.8% | 8.3% |
| F, p | F(3.78,4583)=94.69, p<.001 | F(3.95,4231)=6.69, p<.001 |
| Religion: Catholic | 13.7% | 16.4% |
| Religion: Other Christian | 16.0% | 12.0% |
| Religion: Islam | 47.6% | 15.4% |
| Religion: Traditionalist | 45.8% | 41.3% |
| F, p | F(3.59,4356)=98.09, p<.001 | F(3.39,3626)=2.94, p=.026 |
| Insurance: No | 38.1% | 14.6% |
| Insurance: Yes | 9.8% | 10.6% |
| F, p | F(1,1212)=29.15, p<.001 | F(1,1070)=0.80, p=.372 |
| Facility delivery: Home | 54.0% | 16.2% |
| Facility delivery: Facility | 16.7% | 13.3% |
| F, p | F(1,1209)=399.11, p<.001 | F(1,1067)=3.74, p=.053 |
| ANC 4+: No | 61.2% | 18.6% |
| ANC 4+: Yes | 19.6% | 13.1% |
| F, p | F(1,1201)=509.02, p<.001 | F(1,1058)=9.63, p=.002 |
| Access barrier (any): No | 33.9% | 13.0% |
| Access barrier (any): Yes | 39.9% | 15.7% |
| F, p | F(1,1212)=10.90, p=.001 | F(1,1070)=3.40, p=.065 |
| Media exposure: No | 46.7% | 16.5% |
| Media exposure: Yes | 21.2% | 12.1% |
| F, p | F(1,1212)=159.61, p<.001 | F(1,1070)=8.94, p=.003 |
| Child sex: Male | 38.2% | 13.8% |
| Child sex: Female | 36.4% | 15.1% |
| F, p | F(1,1212)=1.21, p=.272 | F(1,1070)=0.76, p=.384 |

The bivariate pattern for dropout was quite different. Child sex, urban/rural living, ethnicity, and health insurance status were not significantly related to dropout (all p > 0.05) but were strongly related to zero-dose status. Only maternal education (higher education: 6.4% dropout versus 17.1% among mothers with no education; F(2.97, 3182.30) = 7.56, p < 0.001), wealth (richest quintile: 8.3% versus 16.7% among the poorest; F(3.95, 4231.06) = 6.69, p < 0.001), antenatal care (13.1% versus 18.6%; F(1, 1058) = 9.63, p = 0.002), and religion (driven primarily by an elevated 41.3% dropout rate among the small traditionalist subgroup; F(3.39, 3625.68) = 2.94, p = 0.026) retained statistical significance, and effect sizes were consistently smaller than for zero-dose.

### 4.4 Multivariable Logistic Regression

In the fully adjusted model for zero-dose status (Table 4; F(27, 1169) = 26.00, p < 0.001, n = 4,680), maternal education remained independently protective in a clear dose-response pattern (primary: aOR = 0.66, 95% CI 0.49–0.89; secondary: aOR = 0.43, 95% CI 0.32–0.57; higher: aOR = 0.23, 95% CI 0.13–0.41, all p ≤ 0.006), as did facility delivery (aOR = 0.55, 95% CI 0.44–0.69, p < 0.001) and four or more antenatal visits (aOR = 0.33, 95% CI 0.27–0.41, p < 0.001) — the strongest protective factor in the model. After adjustment, only the richest wealth quintile was independently significant (aOR = 0.35, 95% CI 0.21–0.59, p < 0.001), suggesting that the majority of the raw wealth gradient in Table 3 was attributable to the education and facility access gradient. After controlling for zone, ethnicity, wealth, and education, the association between Muslim religious affiliation and increased odds of being a zero-dose child remained statistically significant when compared to Catholic religious affiliation (aOR = 1.67, 95% CI 1.08–2.58, p = 0.022). The odds of having a zero-dose child increased by 11% (aOR = 1.11, 95% CI 1.03–1.20, p = 0.008) for each additional reported access barrier and decreased slightly with each unit increase in birth order (aOR = 0.93 per unit increase in birth order, p = 0.001). Notably, after adjustment there was a sign reversal between education and wealth; facility access and religion; and zonal category (the South West had the second lowest raw zero dose prevalence in Table 3 but the adjusted odds were higher compared to the North West reference category), meaning that for each of these factors, the gap narrowed significantly between the zonal categories and a smaller residual gap remained once the other factors were held constant.

**Table 4.** Multivariable survey-weighted logistic regression: adjusted odds ratios (95% CI), full specification. Gap A: n=4,680, F(27,1169)=26.00, p<.001. Gap B: n=2,982, F(27,1027)=3.33, p<.001. Asterisked (*) values denote p<.05.

| Predictor / Category | Gap A: aOR (95% CI) | Gap B: aOR (95% CI) |
| --- | --- | --- |
| Rural (ref: urban) | 0.78 (0.60–1.01) | **0.44 (0.31–0.61) \*** |
| Zone – North East (ref: NW) | **0.47 (0.34–0.65) \*** | 0.85 (0.54–1.35) |
| Zone – North Central | 0.94 (0.65–1.34) | 1.31 (0.77–2.23) |
| Zone – South East | 0.42 (0.16–1.14) | 1.54 (0.54–4.39) |
| Zone – South South | **0.45 (0.26–0.77) \*** | 0.59 (0.29–1.17) |
| Zone – South West | 1.25 (0.71–2.21) | 0.93 (0.43–2.04) |
| Education – Primary (ref: none) | **0.66 (0.49–0.89) \*** | 0.88 (0.58–1.35) |
| Education – Secondary | **0.43 (0.32–0.57) \*** | 0.87 (0.58–1.31) |
| Education – Higher | **0.23 (0.13–0.41) \*** | **0.45 (0.25–0.81) \*** |
| Ethnicity – Hausa (ref: Fulani) | 0.86 (0.60–1.23) | 0.79 (0.46–1.38) |
| Ethnicity – Igbo | 0.61 (0.25–1.46) | 0.70 (0.25–1.95) |
| Ethnicity – Yoruba | 0.67 (0.36–1.24) | 1.33 (0.61–2.92) |
| Ethnicity – Other | 0.81 (0.57–1.17) | 1.04 (0.61–1.78) |
| Wealth – Poorer (ref: poorest) | 1.02 (0.81–1.30) | 1.33 (0.89–1.99) |
| Wealth – Middle | 0.87 (0.62–1.22) | 1.01 (0.65–1.58) |
| Wealth – Richer | 0.82 (0.57–1.19) | 0.79 (0.48–1.28) |
| Wealth – Richest | **0.35 (0.21–0.59) \*** | **0.46 (0.25–0.82) \*** |
| Religion – Other Christian (ref: Catholic) | 1.15 (0.76–1.74) | 0.75 (0.47–1.20) |
| Religion – Islam | **1.67 (1.08–2.58) \*** | 0.76 (0.44–1.32) |
| Religion – Traditionalist | 2.86 (0.91–8.99) | **3.04 (1.17–7.95) \*** |
| Health insurance (ref: no) | 0.66 (0.24–1.86) | 1.38 (0.58–3.24) |
| Facility delivery (ref: home) | **0.55 (0.44–0.69) \*** | 0.93 (0.70–1.23) |
| ANC 4+ visits (ref: no) | **0.33 (0.27–0.41) \*** | 0.76 (0.56–1.02) |
| Access barrier count (per unit) | **1.11 (1.03–1.20) \*** | **1.15 (1.02–1.29) \*** |
| Birth order (per unit) | **0.93 (0.89–0.97) \*** | 0.98 (0.92–1.05) |
| Female (ref: male) | 0.93 (0.78–1.10) | 1.13 (0.89–1.45) |
| Media exposure weekly+ (ref: none) | 0.94 (0.72–1.22) | 0.92 (0.69–1.23) |

Although there was a strong raw association with media exposure, after adjusting, the effect of media exposure did not differ significantly from the null hypothesis (aOR = 0.94, p = 0.621), suggesting that the unadjusted association is simply mediated by similar socioeconomic and access factors that are already represented in the education, wealth, and facility variables.

The adjusted model for dropout (F(27, 1027) = 3.33, p < 0.001, n = 2,982) had even fewer significant predictors, and several of these predictors changed direction from the zero-dose model. The association of rural residence with lower dropout (aOR = 0.44, 95% CI 0.31–0.61, p < 0.001) was contrary to its (non-significant) association with higher risk of zero doses, indicating that rural children who do not drop out of the schedule may have greater provider continuity than urban children, who may have increased risk of provider fragmentation. The socioeconomic predictors were only protective for higher maternal education (aOR = 0.45, p = 0.008) and the richest wealth quintile (aOR = 0.46, p = 0.008); the graded associations with lower levels of education and wealth observed for zero-dose were not observed for dropout. The unadjusted dropout risk was higher for girls than boys (Table 3); this difference was not significant following adjustment (aOR = 1.13, p = 0.321). There was no independent association between facility delivery, health insurance, and dropout, either positive or negative. We can visualize this difference in predictor profiles between Gap A and Gap B in Figure 2 below, where adjusted odds ratios (and 95% confidence intervals) for a selected subset of important predictors are plotted together for both models.

**Figure 2.**
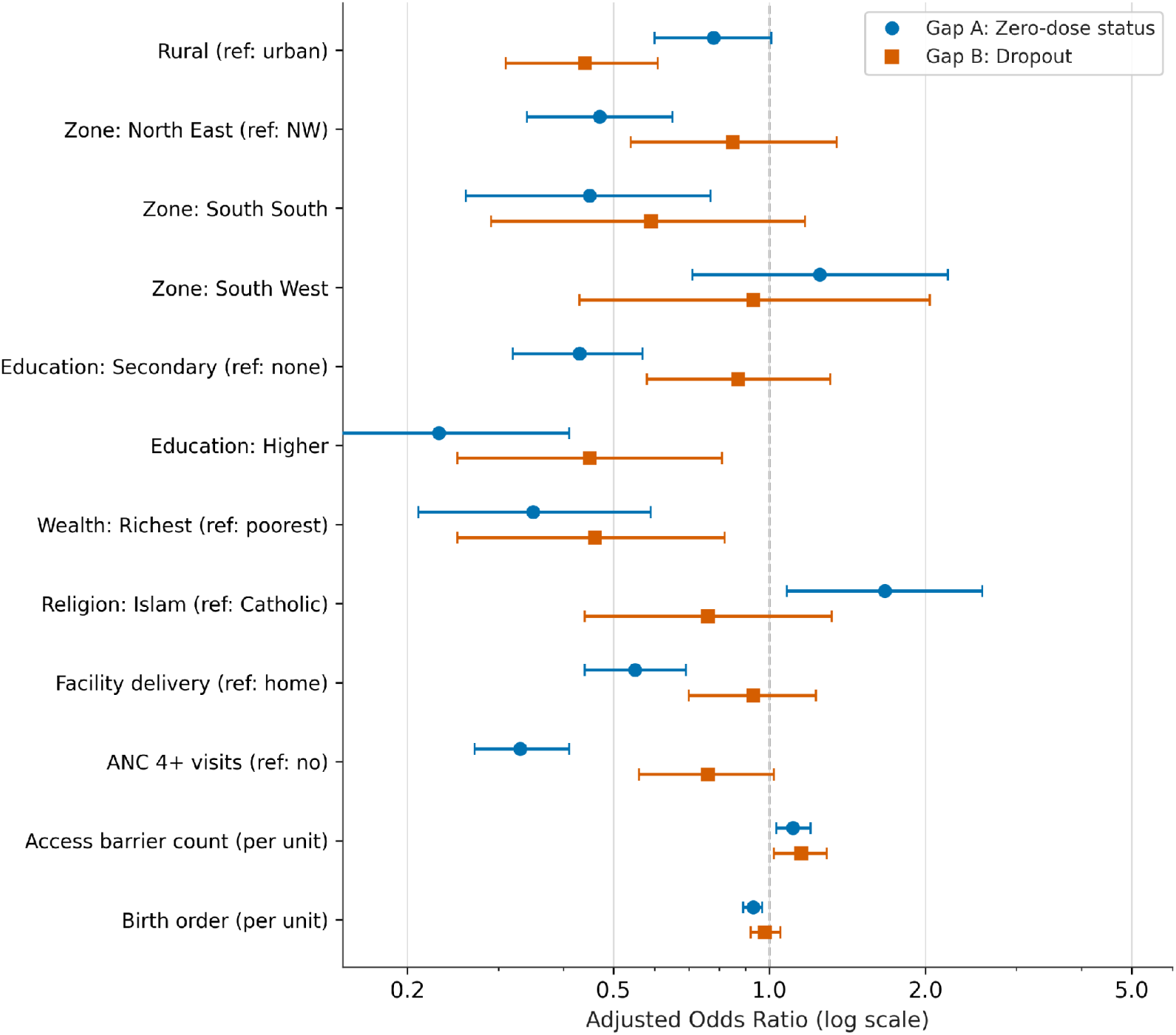
Adjusted odds ratios (95% CI) for key predictors, Gap A (zero-dose) versus Gap B (dropout). Estimates from the fully adjusted survey-weighted logistic regression models (Table 4). Dashed line indicates the null value (OR = 1).

### 4.5 Machine-learning Algorithm Comparison

Table 5 and Figure 3 show the performance results of the algorithms that were not held out as discriminative. The survey-weighted logistic regression method achieved the highest AUC (0.825), with random forest (0.810), elastic net (0.805), and XGBoost (0.794) following. However, the same pattern held for dropout: logistic regression (0.613), random forest (0.594), elastic net (0.584), and XGBoost (0.534), just slightly above random chance. In both cases, the easiest in the comparison set beat all three machine-learning options, and XGBoost was the least effective of the three. The variable importance by random forest was generally similar to the logistic regression results, where antenatal care attendance, maternal education, facility delivery, and wealth were the top four predictors of zero-dose status, and birth order, access barriers, wealth, and education were the top four predictors of dropout, each reflecting the same substantive pattern seen in the regression analysis.

**Figure 3.**
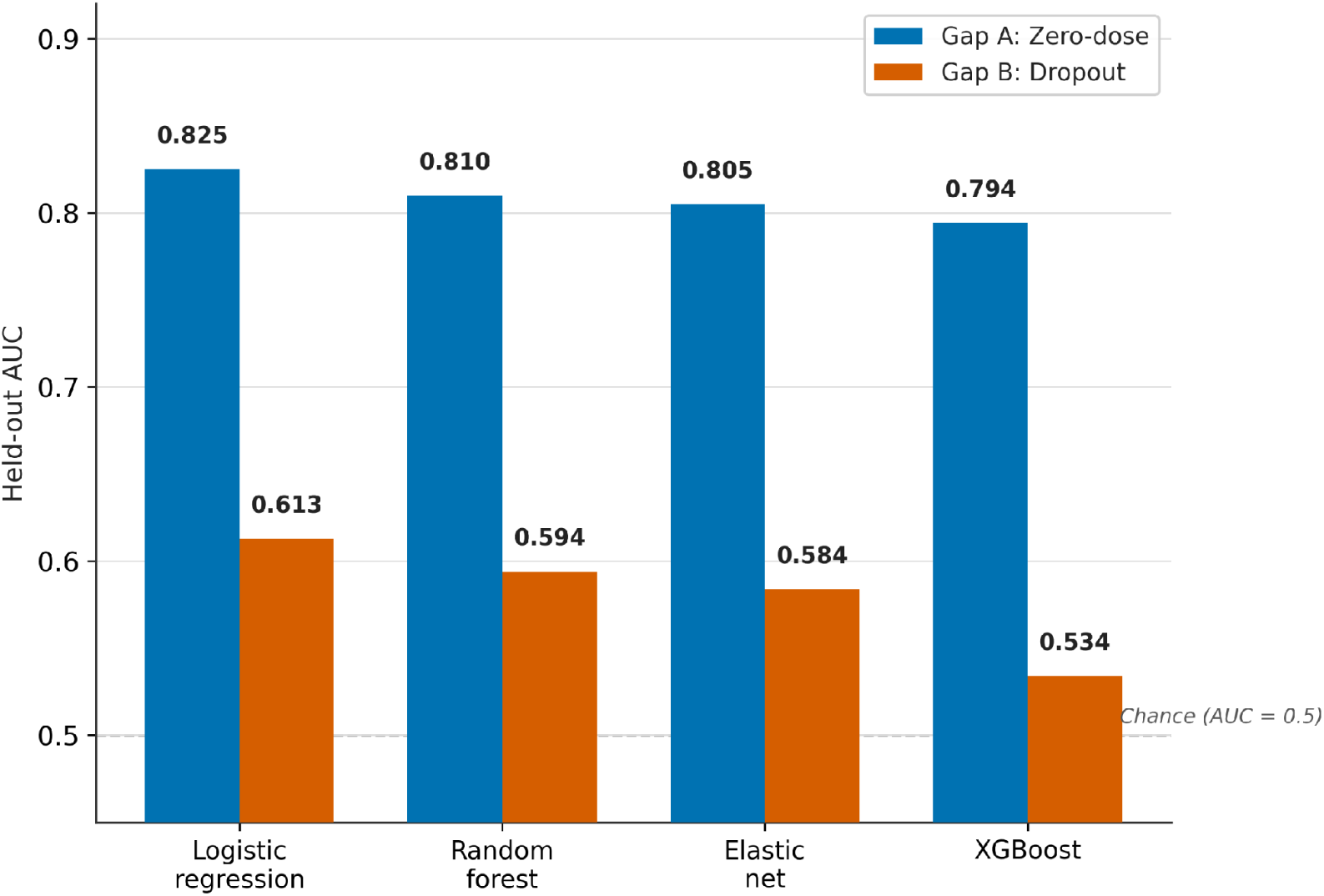
Held-out AUC by algorithm and outcome, primary (12–23 month) sample. Dashed line indicates chance-level discrimination (AUC = 0.5).

**Table 5.**
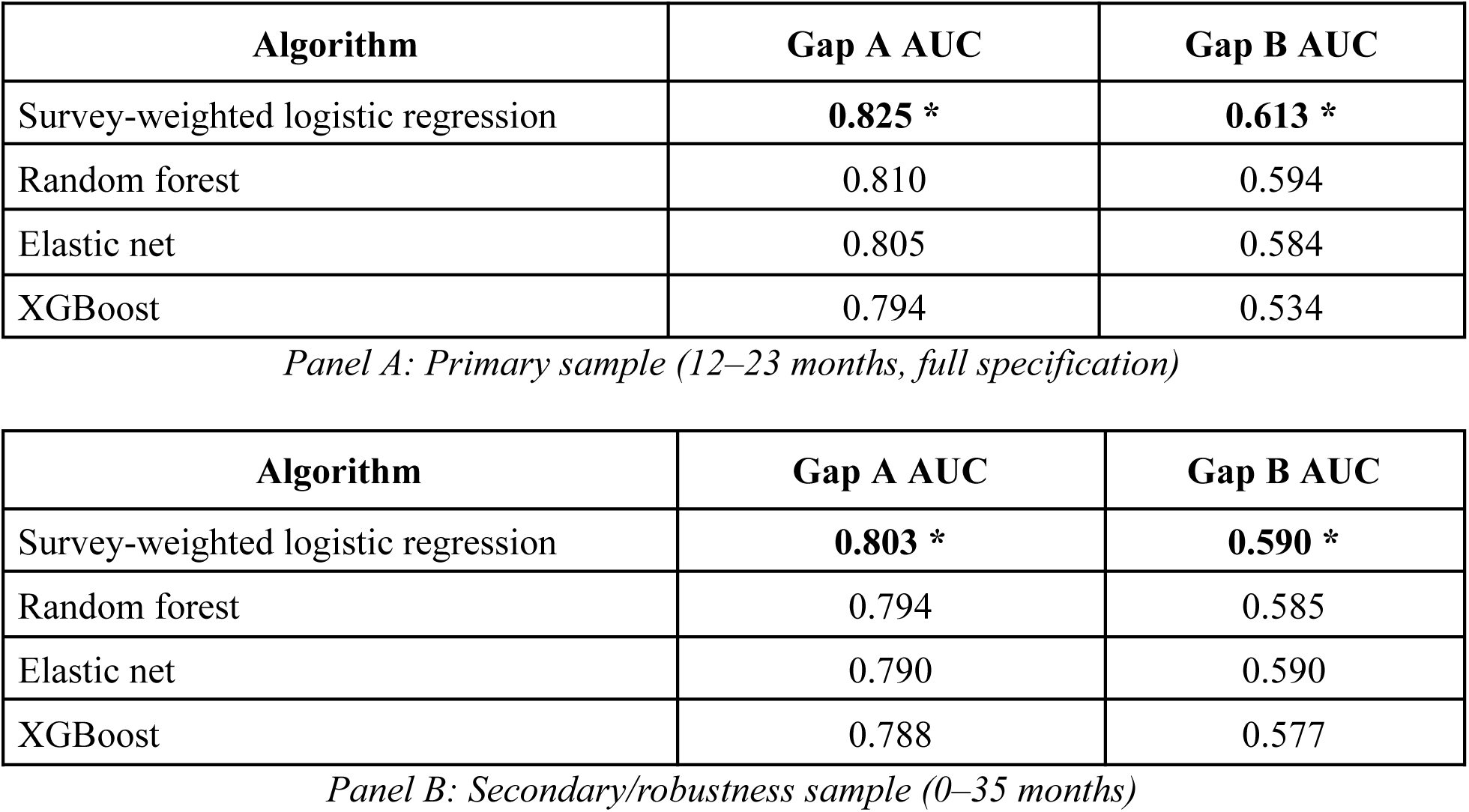
Held-out AUC comparison across four algorithms. Note: Gap A outcome split once (80/20) stratified and seed documented and applied consistently across all algorithms and both samples. Highlighted values with bold/asterisk are top performing algorithm for each outcome/sample.

### 4.6 State-level Geographic Clustering

After adjustment for all individual- and household-level covariates, the intraclass correlation coefficient [ICC] for the random intercept model at the state level was 0.112 (95% CI 0.071–0.172) for zero-dose status and 0.047 (95% CI 0.025–0.085) for dropout status. The fixed-effect estimates of the multilevel model largely confirmed the standard, survey-weighted logistic regression model; most importantly, the association between being Muslim and being zero-dose remained significant (aOR = 1.84, 95% CI 1.31–2.58, p < 0.001) after accounting for state-level clustering, suggesting that the association is not only due to the states where Islam is dominant.

### 4.7 Robustness to Age Restriction

The ordering of the algorithms did not change when the full 0–35 month sample (n = 13,261 for Gap A, n = 7,815 for Gap B) was compared over the same four algorithms, though absolute AUC values were slightly lower. This is consistent with the broader sample’s inclusion of younger infants (under 12 months) who are not yet old enough to have received later vaccine doses, adding noise to the zero-dose classification that the primary 12–23 month sample deliberately avoids.

## 5. Discussion

### 5.1 Principal Findings

Just over half (53.6%) of children 12–23 months in Nigeria are fully vaccinated (pentavalent vaccine, DPT3-HepB-Hib), while more than a third (37.3%) had received none of the vaccine. In addition to the updated national estimates, the main methodological contributions of this study are as follows: (1) the predictors of zero-dose status and dropout were very different, with zero-dose status having more to do with geography and dropout more to do with child and family characteristics; (2) a state-weighted logistic regression model performed best for both outcomes in explaining the residual variation, with geography accounting for more than twice the residual variation in zero-dose status than in dropout status after the model was adjusted; and (3) three more sophisticated machine learning algorithms performed worse than this state-weighted logistic regression model. All of these findings have consequences that extend beyond Nigeria’s immunization program in particular.

### 5.2 Zero-Dose and Dropout Are Different Problems, Not One Problem Measured Twice

There were significant differences between the predictor profiles of the two outcomes. This was clearly reflected in the robust and graded associations with maternal education, socioeconomic status, facility delivery, and antenatal care attendance, as well as the independent and significant association with maternal religious affiliation, which remained strong after accounting for geopolitical zone, ethnicity, and even state-level random effects. The other measures of dropout, in contrast, lost most of their effect sizes after adjustment, and the model’s overall fit was much poorer (F = 3.33 compared to F = 26.00 for zero-dose). In the same way, this asymmetry directly replicates Sato’s (2023) result that the individual contributions of maternal education and household wealth to the decline in zero-dose and under-immunization prevalence in Nigeria during 2003–2018 are substantial, whereas their effects on the decline in dropouts are weak. The fact that two independent ways of analysis – trend data Blinder-Oaxaca decomposition and survey-weighted machine learning based on a single cross-section – arrive at the same substantive answer increases the likelihood that this is a real characteristic of the Nigerian immunization system and not merely due to the nature of either method.

The actual impact is that just trying to reduce the zero-dose prevalence won’t help you reduce dropout, and vice versa. The zero-dose status is a response to the standard determinant that is the target of demand-generation and structural-access interventions: maternal education campaigns, incentives for facility delivery, and linkage to antenatal-care services, which are also contact points in the vaccination system. More meaningful factors underlying dropout that may be included in the predictor set elements are factors that were not well captured by the standard DHS harmonized-control battery such as quality of follow-up by the health worker, continuity of vaccine supplies at the health facility, reminder and defaulter-tracing systems, and the logistics of return visits. This is in line with the international trend of relatively stable dropout rates despite reduction in zero dose prevalence observed in the latest WUENIC release (WHO/UNICEF, 2026).

### 5.3 The Religion Finding and Its Historical Context

Given the number of factors that were controlled, it is difficult to believe that the robust and independent relationship between Muslim religious affiliation and zero-dose status is explained by any of the other factors. One possible contributing factor is Nigeria’s particular vaccine history; the boycott of the oral polio vaccine (OPV) in 2003–2004 by the political and religious leaders of several northern states, spurred by rumors that the vaccine contained anti-fertility agents, was investigated as an example of institutional mistrust expressed through religious leaders rather than a broader anti-vaccine sentiment (Jegede, 2007). Further research on the aftermath of the boycott found that the social norms it produced also continued to thrive within communities and, importantly, beyond the initial information trigger, meaning they became self-reinforcing (Ghinai et al., 2013). A possible but unstudied mechanism in this data is whether this is also true for routine immunization, and this may represent one way that religious affiliation continues to be predictive even 20 years later, and even for a new vaccine series altogether (Leke et al., 2020). The interpretation in this paper should be understood as a possible contributing explanation, but not as a finding directly demonstrated by the data presented here, since the data cannot distinguish between institutional-trust effects and other unmeasured correlates of religious affiliation.

### 5.4 Rural Residence Protects Against Dropout but Not Against Starting

Following this, one of the more surprising findings in this study was that although there was no significant independent risk of not being zero-dose in rural areas, rural residence was associated with a lower risk of dropping out (aOR = 0.44). A potential explanation is when a child has already initiated contact with a health system, the rural health system is more likely to use outreach models and/or more likely to use community health workers, whereas urban families, even though they may have less to gain from initiating contact, could have more health systems to choose among, more opportunities for health systems to change, or less health system involvement with a single system that monitors and calls them back for subsequent doses. This finding can’t be evaluated further with the current data; it makes it clear that dropout-reduction interventions based on urban assumptions (such as digital reminder programs assuming that everyone has a smartphone) may need to be rethought for urban settings specifically; and rural dropout-prevention infrastructure can be relatively effective and may not need to be reworked.

### 5.5 Algorithmic Complexity Did Not Improve Prediction

In both the outcome and both the age specifications tested, the best held-out discriminatory performance was obtained by survey-weighted logistic regression, while XGBoost consistently performed the worst. This is contrary to a commonly held default view of an applied machine learning model that gradient boosting is superior to simpler regression models, but it is consistent with a similarly designed analysis of uncontrolled hypertension based on the ENSANUT survey from Mexico in which XGBoost also underperformed when compared with logistic regression and random forest models (Mendoza-Cano et al., 2025). The exact pattern is not as generalizable as simple beats complex: In the Mexican study, random forest performed better than logistic regression, whereas in this study logistic regression outperformed all three alternatives (random forest). The specific, replicated lesson is that the benefits of XGBoost are not automatically valid for structured, moderate-dimensional sociodemographic survey data, but must be demonstrated in the specific application of interest — a “modest, but useful” result for researchers faced with a similar survey-based prediction problem. It is also interesting to note that even the best-performing model (AUC = 0.61) for dropout had only modest predictive performance, further emphasizing the above interpretation that the standard set of DHS predictors is intrinsically limited for this specific outcome, irrespective of the algorithm used.

### 5.6 Geography Matters More for Zero-Dose Than for Dropout

The state-level intraclass correlation coefficients, which are “independent” lines of evidence for the same underlying distinction of zero-dose and dropout, are 11.2% and 4.7%, respectively. Even when controlling for all predictors possible at the level of the individual and household, the state a child lives in is a significant influence, or in a substantial sense, a place and system phenomenon, when evaluating zero-dose status. Once individual and family circumstances are taken into account, drop-out is fairly evenly distributed across states, indicating that it is more about individual or family circumstances than about what type of health system a family is part of at the state level. This also has implications for resource allocation: it is likely that the higher the proportion of zero-dose children to the total population in a state, and the higher the prevalence in a given geocode, the greater the returns from state-wide, geographically concentrated interventions (such as the Zero-dose programme) versus individualized or facility-level interventions for dropout (in other words, the more concentrated the intervention, the higher the return for Zero-dose, in the North West and North East, versus dropout, which is more individual or facility-specific) (Patenaude et al., 2023).

### 5.7 The Health-Card Exclusion as a Methodological Contribution

Because this study found that health-card retention was not a meaningful predictor when no substantive predictor was provided, it is suggested as a specific methodological warning for future studies that use the DHS immunization modules. In this study, preliminary modeling revealed that adding card retention to the model increased AUC by 0.80 to 0.90, albeit with an implausibly large odds ratio (0.04), which is consistent with cards being a near-restatement of the predicted outcome, as they tend to be issued at the first vaccination contact. In particular, researchers who utilize DHS immunization data for zero-dose prediction should determine whether there is a similar level of risk with card-based variables that are included in their predictor sets, especially when model performance seems to be unusually strong.

### 5.8 Strengths and Limitations

The key strengths of this study are that it uses the recently released nationally representative round of the 2023-24 NDHS rather than an earlier round, it employs a harmonized framework across four analytical approaches (not just one), and the three lines of evidence (bivariate testing, multivariable regression, and multilevel variance decomposition) all point to the same substantive conclusion.

There are several drawbacks to mention. First, this is a cross-sectional analysis and, as such, cannot infer causal relationships: associations between, for example, facility delivery and zero-dose status are likely due to the health system’s causal impact and to residual confounding from unmeasured maternal health-seeking orientation. Second, all vaccination status data are based on maternal recall and/or card documentation, which carries the risk of recall bias and, for the small “don’t know” group, genuine uncertainty, and is not addressed by the sensitivity analysis of the headline zero-dose estimate. Third, some subgroup estimates have broad confidence intervals, such as the category of traditionalist religion (n < 100 weighted) and should therefore be used with care. Fourth, the interlevel weighting in the multilevel model does not account for all levels of clustering and strata, and the intraclass correlation estimates should be interpreted as indicative rather than design-consistent. Finally, results are restricted to a 12–23-month age range; although the same algorithmic ordering was found in the secondary (0–35-month) analysis, some point estimates may not be representative of children outside this age range. Last, while methodologically appropriate, the current study cannot comment on whether this exclusion of health-card retention has an impact on its value as a method, since this could be a legitimate and less circular predictor of the outcome of interest, specifically within the dropout model, where the leakage concern might be structurally less severe than for zero-dose children; this is a question that needs further investigation with data that allow a clearer temporal separation between the health-card and the outcome of interest.

## 6. Conclusion

The findings of this study offer new, nationally representative data indicating that zero-dose status and dropout are two distinct issues in Nigeria’s childhood immunization system, for which different factors predict them, different levels of geographic clustering are observed, and different levels of predictability from standard household-survey data exist. After controlling for these factors, “zero dose” status is significantly and independently related to maternal education, household wealth, facility delivery, attendance at antenatal care, and religious affiliation, and is significantly influenced by geographical location. By contrast, dropout is more difficult to predict from the same factors, indicating that factors behind it are more likely to lie in the continuity of the health system, such as vaccine stock reliability, follow-up system and defaulter-tracking, and logistics of return visits, than in the sociodemographic factors that underpin the zero-dose model. Accordingly, policies that aim at closing the “coverage gap” as a whole may misallocate resources, as the interventions necessary to address the zero-dose gap have a strong geographic and North Western/Northern orientation, but the interventions required to address the dropout gap do not necessarily have a geographical orientation and are not clearly related to household characteristics.The findings of this study offer new, nationally representative data indicating that zero-dose status and dropout are two distinct issues in Nigeria’s childhood immunization system, for which different factors predict them, different levels of geographic clustering are observed, and different levels of predictability from standard household-survey data exist. After controlling for these factors, “zero dose” status is significantly and independently related to maternal education, household wealth, facility delivery, attendance at antenatal care, and religious affiliation, and is significantly influenced by geographical location. By contrast, dropout is more difficult to predict from the same factors, indicating that factors behind it are more likely to lie in the continuity of the health system, such as vaccine stock reliability, follow-up system and defaulter-tracking, and logistics of return visits, than in the sociodemographic factors that underpin the zero-dose model. Accordingly, policies that aim at closing the “coverage gap” as a whole may misallocate resources, as the interventions necessary to address the zero-dose gap have a strong geographic and North Western/Northern orientation, but the interventions required to address the dropout gap do not necessarily have a geographical orientation and are not clearly related to household characteristics.

Methodologically, the study did not reveal any benefit of algorithmic complexity; survey-weighted logistic regression had the best performance for both outcomes, with the latter being the worst, a result that mirrors a similar finding in an independent study of hypertension in Mexico, albeit in a different context. The superiority of complex machine learning approaches should be tested as a hypothesis rather than assumed to inform a study design in problems that are similarly structured and moderately dimensional in the survey-prediction context.

Future studies need to focus on data collection that captures health-system-level drivers of dropout that the predictor set used in this study could not, and on longitudinal or facility-linked designs that can speak to causal mechanisms that can only be described in this cross-sectional study.

## Declarations

### Conflict of Interests

N/A

### Ethics Approval and Consent to Participate

The data for this study are secondary data and come from de-identified, publicly available data from the 2023–24 Nigeria Demographic and Health Survey (NDHS). The original survey team obtained informed consent from all respondents, and the original NDHS protocol was approved by the National Health Research Ethics Committee (NHREC) of Nigeria and the ICF Institutional Review Board. As this analysis used only de-identified secondary data obtained through registered, authorized access to the DHS Program repository, it was exempt from additional institutional review.

### Clinical Trial Registration

N/A

### Funding Sources

N/A

### Artificial Intelligence Statement

This work is not generated by a Generative Artificial Intelligence (G.A.I.) or large language model (L.L.M.) tool. The information provided is the authors’ own views and opinions.

## Acknowledgements

The authors are thankful to Ahead Labs, IIT Roorkee, for providing Stata 15 and the necessary skillset necessary for this project. The authors also acknowledge Marwadi University for providing the resources that helped in the successful conduct of the research.

## Data Availability Statement

The datasets analyzed in this study are available from the DHS Program (https://dhsprogram.com) but are not publicly available due to restrictions protecting respondent confidentiality. Access requires registration and approval from the DHS Program and is granted upon reasonable request at https://dhsprogram.com/data/available-datasets.cfm. The specific dataset used was the 2023–24 Nigeria Demographic and Health Survey (NDHS) Children’s Recode file. Other literature and additional information was extracted from publicly accessible clinical trial data and World Health Organization (WHO) reports.

## Large-Language Model (LLM)

N/A

## Notes

### Competing Interest Statement

The authors have declared no competing interest.

### Author Declarations

The 2023-24 Nigeria Demographic and Health Survey (NDHS). Link: https://dhsprogram.com/data/dataset/Nigeria_Standard-DHS_2024.cfm?flag=1

